# Prevalence of Methicillin-Resistant *Staphylococcus aureus* in Diabetic Patients Visiting the 37-MilitaryHospital in the Accra Metropolis

**DOI:** 10.1101/2023.11.27.23299071

**Authors:** Akosua Serwaa Boaitey-Biko, Ishmael Bukari, Isaac Ogbe, Michael Acheampong Debrah

## Abstract

Misuse of antibiotics contributes to the worldwide rise in antibiotic resistance. However, it is equally important to consider inevitable situations that may contribute to a rise in antibiotic resistance and implement best practices to ensure the minimality of their resulting adverse effects. A case in point is the increasing prevalence of methicillin-resistant *Staphylococcus aureus* (MRSA), a strain of bacterium that has developed resistance to the antibiotic methicillin and other lactams among long-term diabetic patients as observed in this study. Diabetes is associated with immunosuppression and foot ulcers, which require the constant use of antibiotics as prophylactics, as such, persons with this condition would hold bacterial populations subject to immense selective pressure and are therefore at increased risk for MRSA colonization. Furthermore, these individuals may serve as reservoirs and disseminate these highly drug-resistant strains on return to the general populace. This warranted an investigation into the prevalence of MRSA among diabetic patients at the 37-Military Hospital. A total of 50 samples were taken from the sampled Military Hospital by swabbing the participants’ skin, nose, and wounds aseptically, and 20 samples were acquired from non-diabetics to serve as controls. To culture the samples, blood, and mannitol salt agar were used. Bacteria were isolated and identified using Gram staining and other biochemical testing. The Kirby-Bauer disc diffusion method was used with Muller-Hinton agar to test for antimicrobial susceptibility. According to the study’s findings, antibiotic resistance was much higher among diabetic patients than among non-diabetic patients at the 37-Military Hospital in the Accra Metropolis, Ghana.

## Introduction

The incidence of diabetes mellitus (DM), a chronic metabolic disorder characterized by elevated blood sugar levels, has reached epidemic proportions worldwide including Ghana (1,2). The presence of diabetes lowers the immune system, which increases the body’s susceptibility to infections (3). Among people with diabetes, methicillin-resistant *Staphylococcus aureus* (MRSA) is a leading cause of various infections, including skin, soft tissue, the urinary tract, heart, and blood vessels (4–6) MRSA is a distinctly evolved strain of the bacterium *Staphylococcus aureus* that has developed resistance to multiple antibiotics, including methicillin, beta-lactam antibiotics, and cephalosporins, greatly increasing the recalcitrance of their associated infections (7–9). Also, studies have shown that diabetic individuals are more susceptible to MRSA colonization and infection than other patient populations during hospital admission (10–14).

Except in cases of severely advanced DM or following a surgical procedure, in Ghana, most diabetic cases are handled as outpatient; where individuals visit hospital clinics for tests and treatment without a need for a hospital stay. This method of patient management without proper monitoring mechanisms would not only put diabetic patients at increased risk of infection leading to complications but also position diabetic individuals as vectors for these drug-resistant pathogens into the general populace on return to their homes and daily activities.

Thus, it is essential to gain an understanding of the incidence of MRSA in the diabetic patient population at major hospitals as well as the risk factors associated with the infection, to direct infection control efforts and improve patient outcomes in Ghana. This study focuses on the 37-Military Hospital, and aims to contribute significantly toward advancing patient care, the creation of targeted interventions, enhanced infection control strategies, and appropriate antibiotic prescribing practices, all of which are intended to minimize the spread of MRSA infections and their impact on this vulnerable group within the Accra Metropolitan Area.

## Results

### Demographic Characteristics

According to the results of the study, the average age of female diabetic patients was somewhat higher than that of male diabetic patients, with a range of 46-82 years for the former and 45-77 years for the latter. We also discovered that the median age of diabetic patients was 54 years old for men and 63 years old for women. In addition, the 3 infants’ ages ranged from 2 to 6, with the average being 4 years.

### Bacterial Profile of Study Participants and Control

Tables 1 & 2 show that *S. aureus* was the predominant bacterium isolated from both groups. We also found that the highest numbers of *S. aureus* isolates were found on the skin of diabetic patients, *S. aureus* was isolated from three different sites: the nose (30%), wounds (5%), and skin (65%) indicating that the skin might serve as a significant reservoir for this bacterium in diabetic individuals.

**Table 1:**
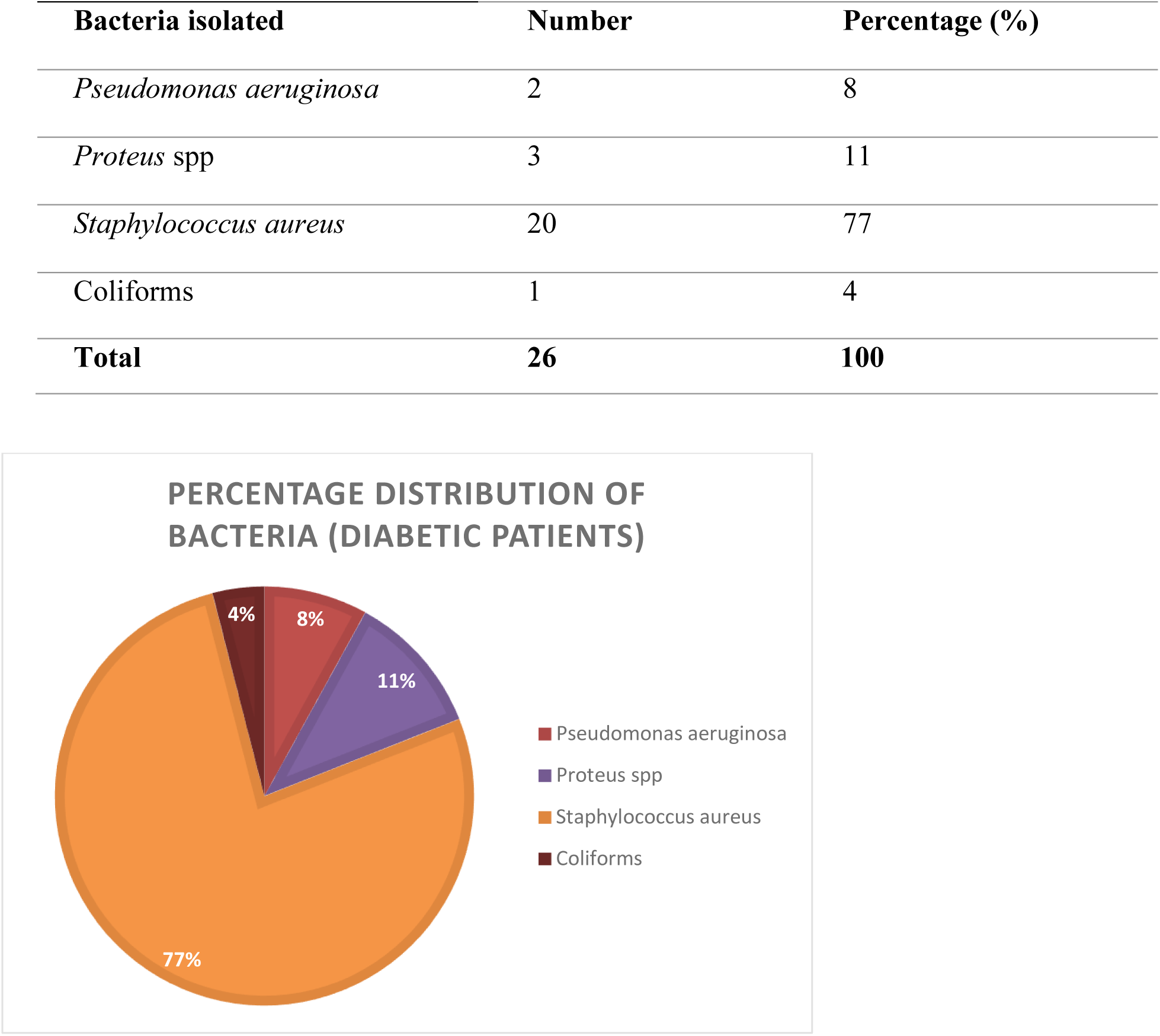
Percentage distribution of bacteria isolated from the diabetic patients selected for the study.

**Table 2:** Percentage distribution of the bacteria isolated from the healthy participants (controls)

| Bacteria isolates | Number of isolates | Percentage (%) |
| --- | --- | --- |
| Staphylococcus aureus | 6 | 86 |
| Pseudomonas aeruginosa | 1 | 14 |
| Total | 7 | 100 |

**Table 3:** Percentage distribution of *S. aureus* isolated from the various body sites of the study participants in 37-Military Hospital, Accra.

| Parts of the body | Diabetic patients | percentage % | Control samples | % |
| --- | --- | --- | --- | --- |
| Nose | 6 | 30 | 2 | 33 |
| Wound | 1 | 5 | 4 | 67 |
| Skin | 13 | 65 |  |  |
| Total | 20 | 100 | 6 | 100 |

In the non-diabetic control group, *S. aureus* was found in the nose (33%) and wounds (67%), with no isolates detected from the skin. This suggests that *S. aureus* may be present in the nasal passages and wounds of non-diabetic individuals, but not as prevalent on their skin compared to diabetic patients. In addition, it was revealed that *Pseudomonas aeruginosa* was present in both groups but at a lower percentage in diabetic patients (8%) compared to non-diabetic controls (14%).

### Differential Statistics between the Isolated *S. Aureus*, Age, Gender, and the Part of the Body from Which Sample Was Collected

From the data presented in Table 4, it is evident that there is a significant association between the prevalence of *S. aureus* and the age of the participants, with a p-value of 0.001. This suggests that the prevalence of *S. aureus* varies with different age groups, highlighting a potential age-related pattern in its occurrence. However, the association between *S. aureus* prevalence and gender was not significant, as evidenced by the p-value of 0.686. This means that there is no significant difference in the prevalence of *S. aureus* between males and females in the study population.

**Table 4:**
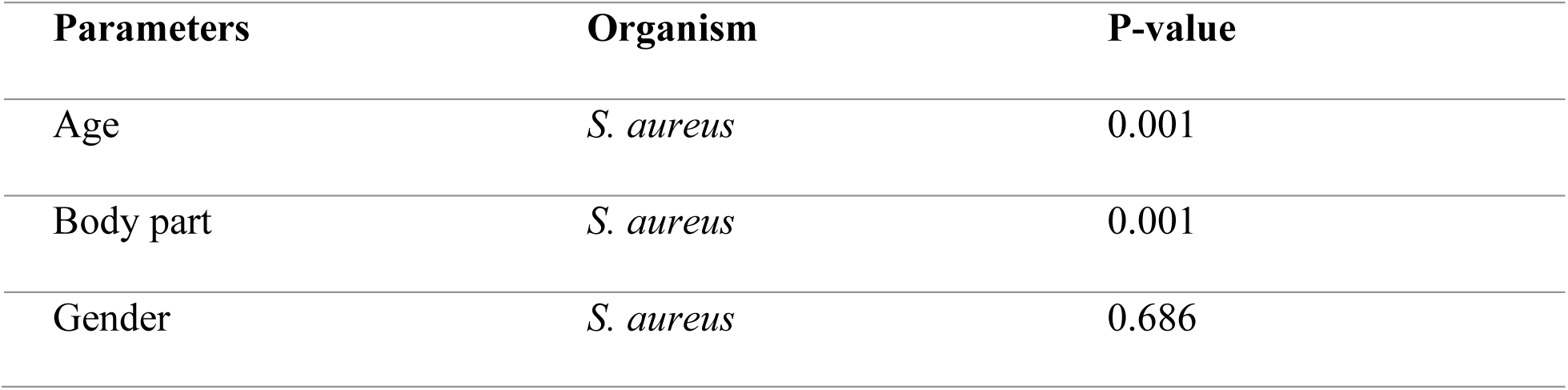
Association between the isolated *S. aureus*, age, gender, and the part of the body from which the sample was collected.

| Parameters | Organism | P-value |
| --- | --- | --- |
| Age | *S. aureus* | 0.001 |
| Body part | *S. aureus* | 0.001 |
| Gender | *S. aureus* | 0.686 |

Another important finding is the significant association between the isolated *S. aureus* and the part of the body from which the sample was collected, with a p-value of 0.001. This indicates that S. aureus colonization differs depending on the location of the sample. In particular, the higher prevalence of *S. aureus* on the skin of diabetic patients may be reflected in this association.

### Percentage Distribution of Isolated MRSA

Figures 1 and 2 highlight that MRSA is more dominant than sensitive *S. aureus* within diabetic patients, for which an opposite trend is observed in non-diabetic patients. Furthermore, in the diabetic patient group, all 6 cases (100%) of *S. aureus* isolated from the nose were identified as MRSA. Additionally, of the 16 MRSA cases among diabetic patients, the majority, 10 cases (62.5%), were isolated from the skin, while 6 cases (37.5%) were isolated from the nose. Notably, no MRSA was obtained from the wounds of diabetic participants. This data highlights that MRSA is particularly common in the nasal passages and skin of diabetic patients.

**Figure 1:**
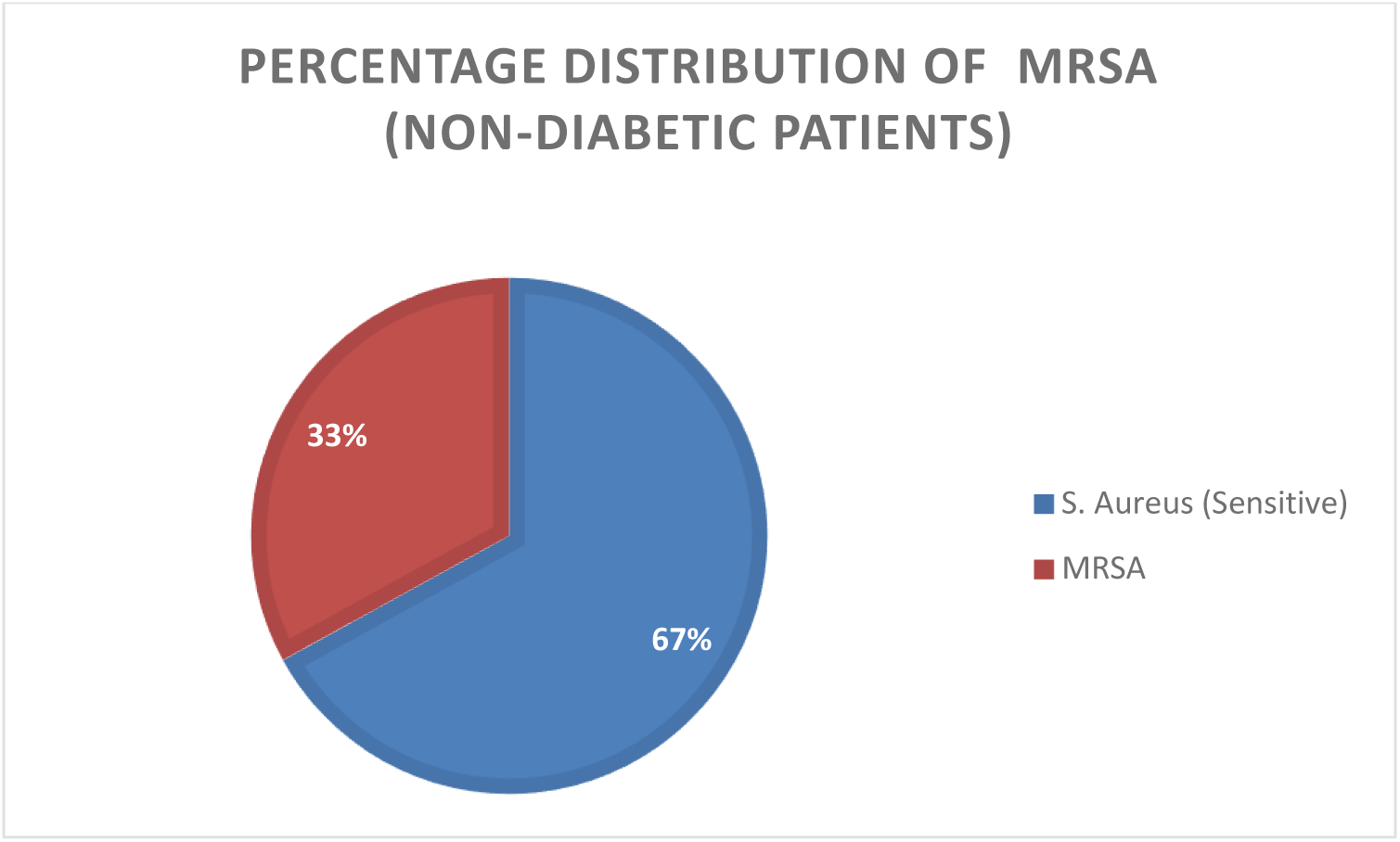
Percentage Distribution of S. aureus and MRSA in non-diabetic patients.

**Figure 2:**
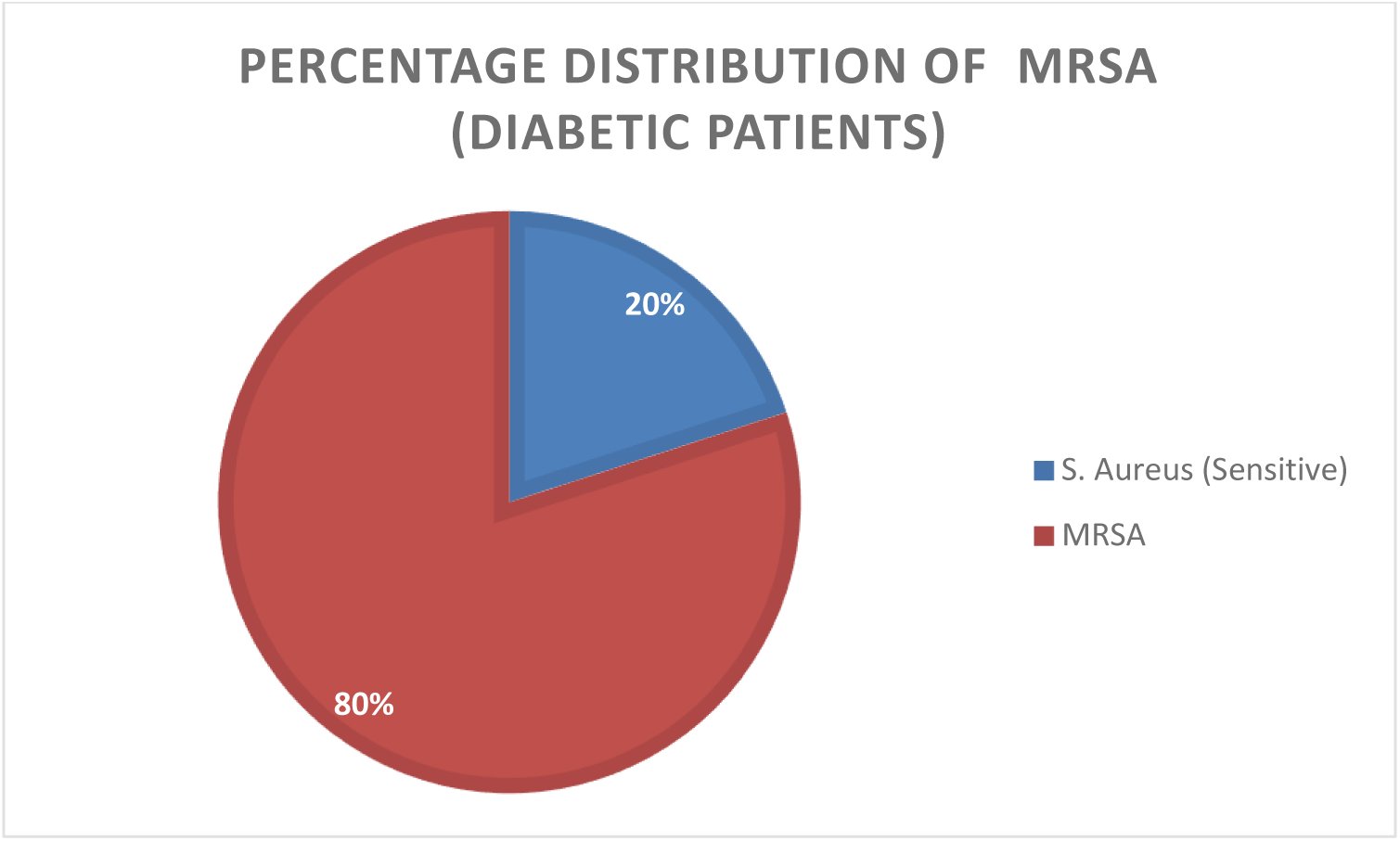
Percentage Distribution of S. aureus and MRSA in diabetic patients.

In the control group, 2 out of 6 cases (33%) of *S. aureus* isolated from the nose were identified as MRSA. Similarly, 1 case (16.5%) each was isolated from the wound and nose of participants in the control group were MRSA.

### Percentage Distribution of Antibiotic Susceptibility Profile of Isolated S. Aureus

In the diabetic patient group, the *S. aureus* isolates showed high resistance to penicillin (100%), erythromycin (85%), and trimethoprim-sulfamethoxazole (85%). They also displayed considerable resistance to cefoxitin (80%), gentamicin (60%), and clindamycin (60%). However, there was relatively low resistance to vancomycin (30%) and tetracycline (20%). The most effective antibiotic was linezolid, with a high sensitivity rate of 85%, and no resistance was observed against it. There were also 4 cases (20%) of intermediate susceptibility to cefoxitin among the *S. aureus* isolates from diabetic patients.

In the non-diabetic control group, *S. aureus* isolates showed high resistance to penicillin (100%) and tetracycline (67%) and moderate resistance to trimethoprim-sulfamethoxazole (50%). However, the highest sensitivity was observed for linezolid, with all isolates showing susceptibility (100%).

Overall, these results indicate that the isolated *S. aureus* strains from diabetic patients exhibited significant resistance to multiple commonly used antibiotics, posing challenges in treating infections caused by these strains. Linezolid appeared to be the most effective antibiotic for diabetic patients and non-diabetic controls. This information is provided in Figures 3 and 4, together with Table 5, showing the breakpoints for antibiotic susceptibility testing, interpreting the results in the context of standard guidelines and assist clinicians in making informed decisions about appropriate antibiotic therapy for MRSA infections.

**Figure 3:**
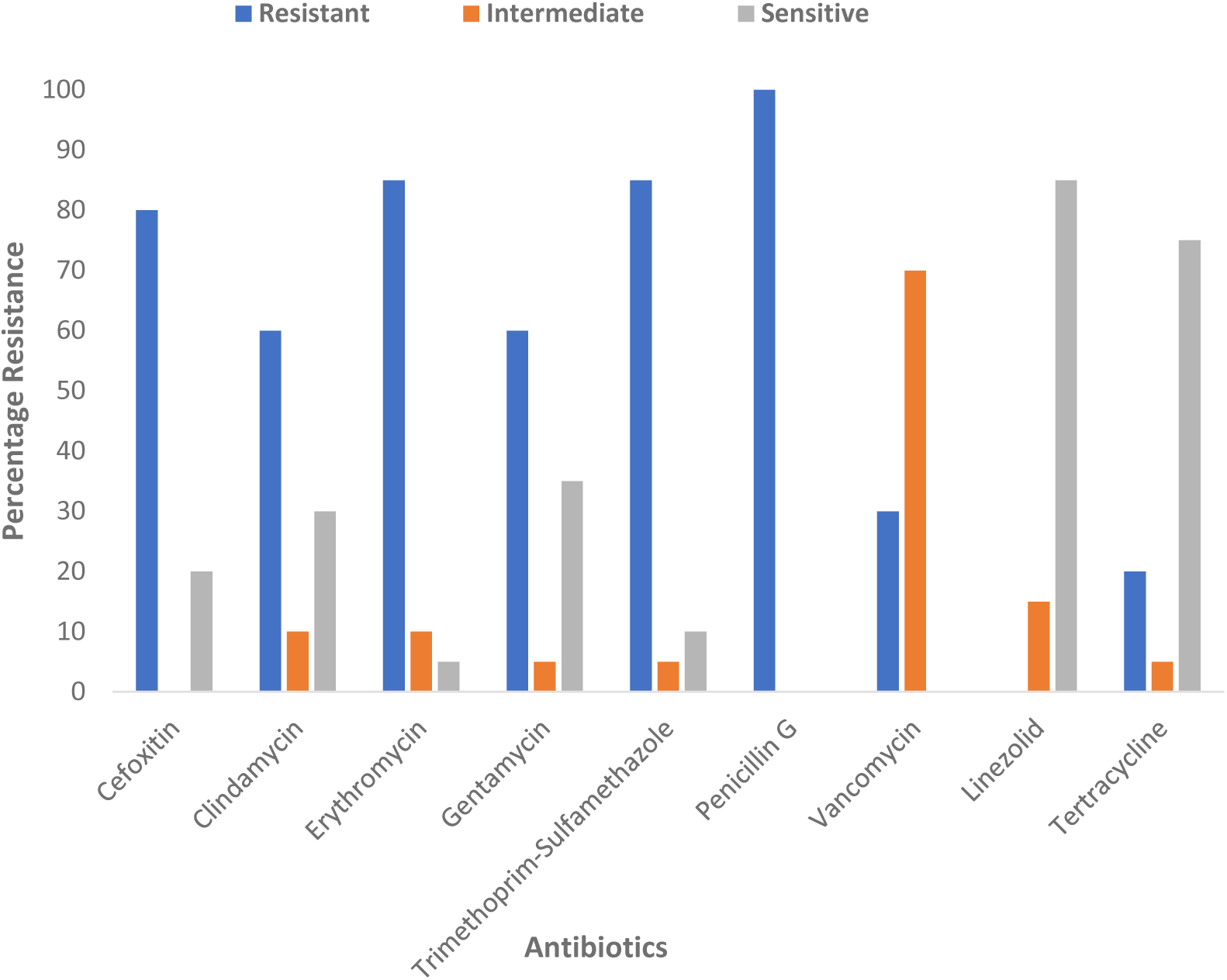
Antibiogram of isolated S. aureus from diabetic parents

**Figure 4:**
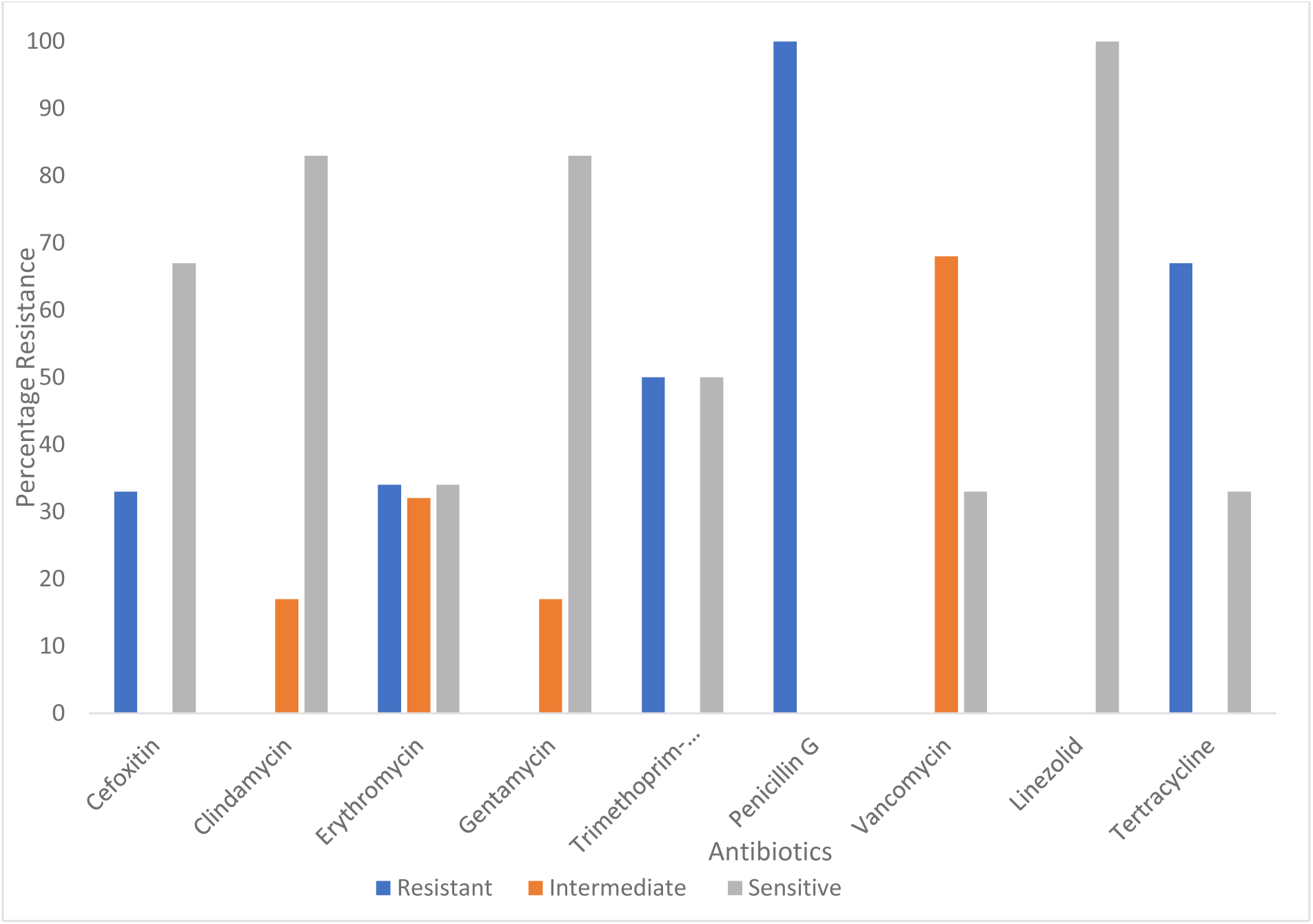
Antibiogram of isolated S. aureus from control samples

**Table 5:** The breakpoints of the antibiotic susceptibility profile as per CLSI 2017.

| Antibiotic |  | Disc content | Zone Diameter |  |  |
| --- | --- | --- | --- | --- | --- |
| Class | Type | (µg) | S≥ | I | R≤ |
| Penicillin | Penicillin G | 1 | 29 |  | 28 |
| Glycopeptide | Vancomycin | 5 | 14 | 11-13 | 10 |
| Aminoglycosides | Gentamycin | 10 | 15 | 13-14 | 12 |
| Macrolides | Erythromycin | 15 | 23 | 14-22 | 13 |
| Tetracyclines | Tetracycline | 30 | 19 | 15-18 | 14 |
| Lincosamides | Clindamycin | 2 | 21 | 15-20 | 14 |
| Folate Pathway Inhibitor | Trimethoprim- Sulfamethoxazole | 25 | 16 | 11-15 | 10 |
| Oxazolidinones | Linezolid | 10 | 21 |  | 20 |
| Penicillinase-Stable Penicillins | Cefoxitin | 30 | 22 |  | 21 |

### Percentage Distribution of Antibiotic Susceptibility Profile of Isolated MRSA

From the data presented in Figures 5 and 6, along with the information in Table 5, within the diabetic patient group, the MRSA isolates exhibited high resistance to penicillin (100%) and trimethoprim-sulfamethoxazole (100%). They also showed considerable resistance to erythromycin (93.75%) and gentamicin (75%). However, there was a relatively low level of resistance to linezolid (6.25%), making it one of the most effective antibiotics against MRSA in this group. Interestingly, there were 11 cases (68.75%) of MRSA isolates with intermediate sensitivity to vancomycin, suggesting some degree of reduced susceptibility to this last-resort antibiotic.

**Figure 5:**
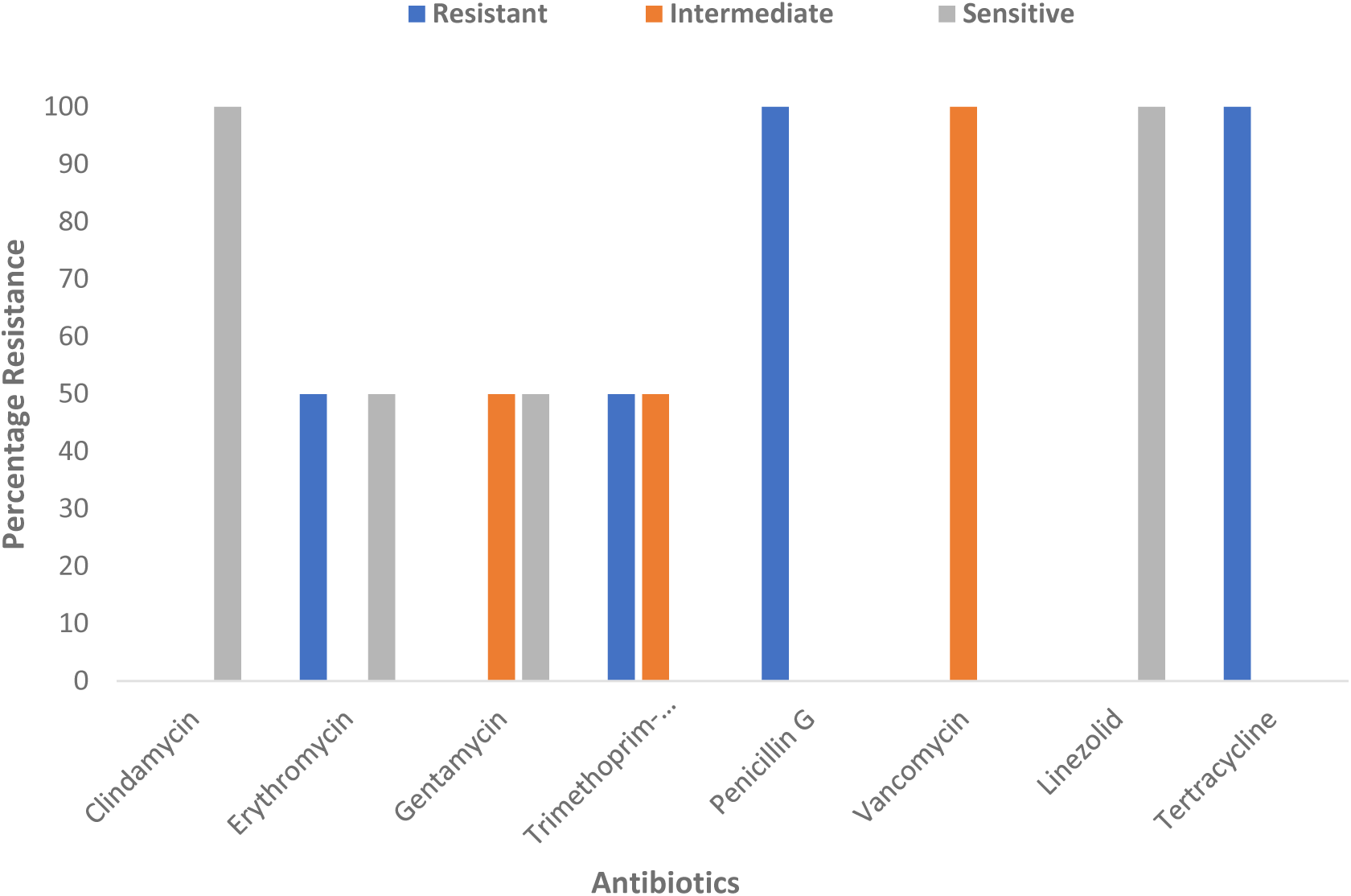
Antibiogram of isolated MRSA from the control sample

**Figure 6:**
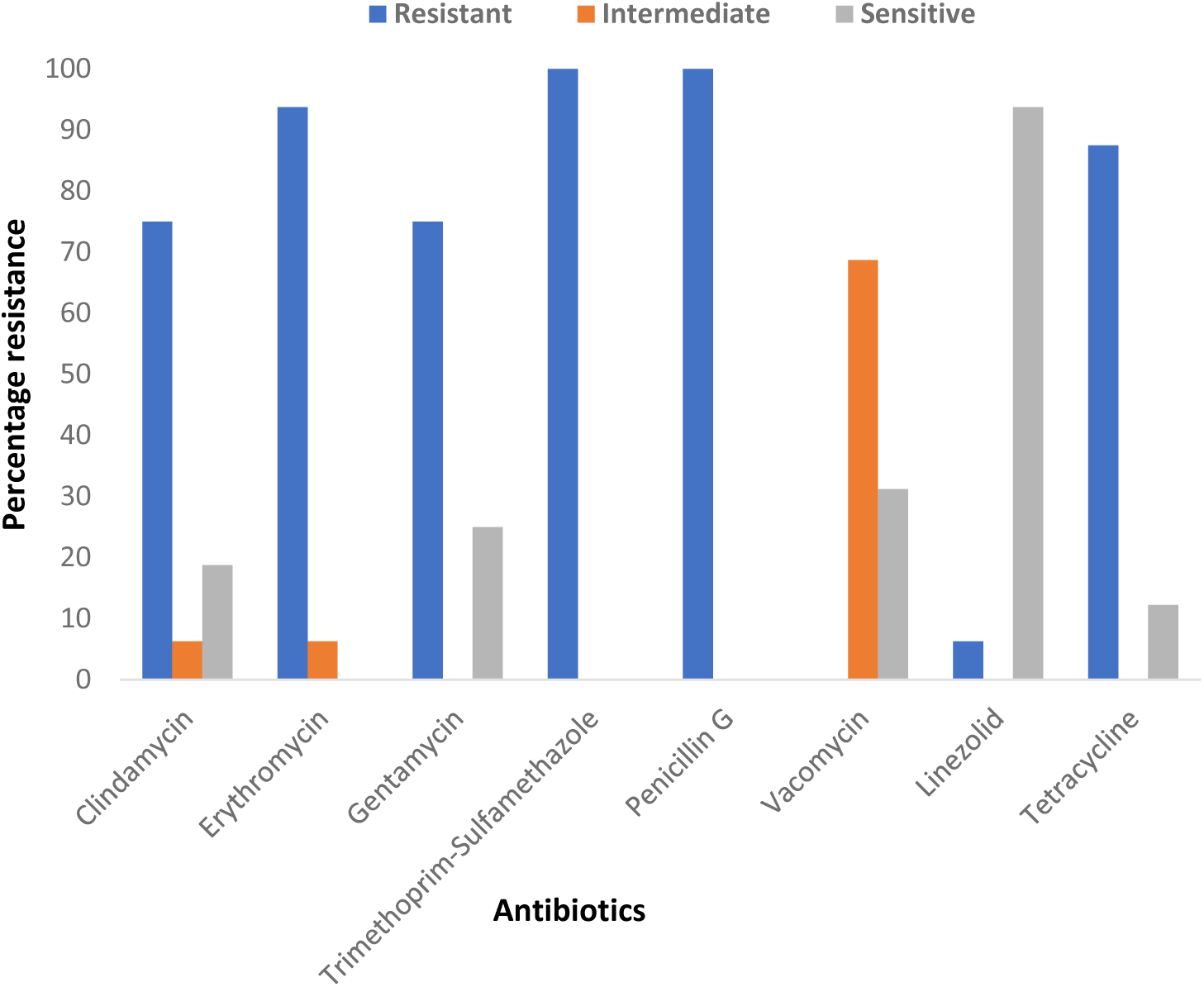
Antibiogram of isolated MRSA from diabetic patients

Overall, the data indicates significant antibiotic resistance among the MRSA isolates from diabetic patients and non-diabetic controls.

## Discussion

From the results, we highlight that the increasing prevalence of MRSA potentially represents a strain-level variation which has been shown in research by Kalan *et al* to be associated with poor clinical outcomes in diabetic patients (15). Although the MRSA prevalence is more prominent in diabetic patients, the lack of competing bacteria within the non-diabetic patients presents a significant risk of a potential epidemic. These findings emphasize the importance of monitoring and controlling MRSA colonization in diabetic patients to reduce the risk of potential infections and improve overall patient outcomes.

Also, the presence of Proteus spp and Coliform which were exclusively isolated from the diabetic patient group are of interest as previous research by Kalan *et al* has shown that non-pathogenic bacteria may contribute to accelerated wound healing but in some other cases may contribute to exacerbated inflammation through the presentation of superantigens and non-specific stimulation of T-cell populations (15). Further analysis of these bacterial isolates may provide valuable insights into their role in clinical outcomes for diabetic patients and non-diabetic controls, potentially aiding in understanding the link between bacterial infections and diabetes.

Looking at the antibiograms of the MRSA strains, an observable interesting pattern is that despite the uniformity displayed by the strains in their extreme sensitivity to linezolid as well as resistance to penicillin and tetracycline across both patient categories (control and diabetic) a significant increase in resistance is presented in specific antibiotics (Clindamycin, Erythromycin, Gentamycin and Trimethoprim-sulfamethoxazole) within the diabetic patient group as compared to the control group. A study by Anafo, R. B., *et al* in 2021 reported similar resistance levels for penicillin (91%-95%) but generally lower resistance levels for other antibiotics (16). Also, the sensitivity to linezolid is reflected in their results suggesting that the significant increase in resistance within the diabetic patient group as compared to the control group in our results may represent a reflection of an environment supporting increased evolutionary pressure driving the generation of highly multi-drug resistant strains which may go beyond drug pressure selection. Identifying any other potential contributors if present would be critical to controlling the emergence of epidemic strains in the future.

The presence of a pre-existing resistant population or a significant intermediately resistant population within these specific antibiotics displaying an increased resistance profile as you move from the control group to the diabetic group lends weight to a transitional theory in which sensitive strains become intermediately resistant before acquiring full resistance. However, the profile of clindamycin appears to deviate from this trend in that, although no intermediately resistant population is observed within the control group, a significant population of highly resistant strains is observed in the diabetic patient group. Similarly, by convention of a transitional theory to the generation of a highly resistant population, we would expect a significant percentage of strains responding in a resistant manner to vancomycin within the diabetic patient group since all the strains tested within the control group were intermediate-resistant. The results, however, show that a percentage of the strains within the diabetic group are sensitive. These sensitive strains however may highlight that a shift to intermediate resistance across strains may not be 100% complete thus far. The presence of vancomycin-intermediate Staphylococcus aureus (VISA) has also been reported in over 50 cases with an overall 1.7% prevalence among approximately 23,000 strains (17). Since vancomycin has been the go-to antibiotic for treating MRSA infections for decades, this and other research suggest that this rising resistance is a problem (17,18). Nevertheless, the distribution of intermediately resistant MRSA to vancomycin indicates a need for close monitoring and control to prevent a shift toward high resistance.

A similar pattern is observed across the antibiograms of *S. aureus* in the control and diabetic patient groups. An increasing pattern of resistance as you transition from the control group to the diabetic group in the same manner across the same specific antibiotics (Clindamycin, Erythromycin, Gentamycin, and Trimethoprim-sulfamethoxazole) as in the case of the MRSA strains. These similarities highlight that factors contributing to the generation of resistant strains within diabetic patients may act uniformly across bacterial strains present.

Overall, the data points to a growing prevalence of highly resistant strains within diabetic patient groups as such, strategies including constant drug resistance surveillance, and antibiotic prescription control are needed if the current outpatient system of handling diabetic patients is to be maintained.

## Materials and Methods

### Introduction

This study investigated the bacterial profile and antimicrobial susceptibility of samples collected from diabetic patients and non-diabetic controls at the 37-Military Hospital. A total of 50 samples were aseptically collected from diabetic patients, including swabs from their skin, nostrils, and wounds. Additionally, 20 samples were obtained from non-diabetic individuals to serve as controls. The samples were cultured on blood and mannitol salt agar, and subsequent isolation and identification of bacteria were performed using Gram staining and other biochemical tests. The susceptibility of the isolated bacteria to antimicrobial agents was assessed using the disk diffusion method on Muller-Hinton agar.

### Sample Collection

Participants for this study were recruited from the diabetic patient population at the 37-Military Hospital across several hospital sites: the Medical and Surgical Outpatient Department (SOPD), Female and Male Medical Ward, Surgical and Polyclinic Dressing Room, and Emergency Room. Written informed consent was obtained from each participant before sample collection. Aseptic techniques were employed during the collection of samples to minimize contamination. For diabetic patients, swabs were taken from their skin, nostrils, and any existing wounds. For the non-diabetic controls, swabs were taken from the skin and nostrils. The cotton tips of the swab sticks were broken into pre-labeled bijou bottles containing 10 ml sterile peptone water and gently re-capped. The collected samples were carefully labeled and transported to the Microbiology Laboratory, School of Biomedical and Allied Health Sciences, College of Health Sciences, University of Ghana, Korle-Bu, for analysis.

### Analysis of the Samples

All the media utilized in this investigation were sterilized (autoclaved at 121C for 15 minutes) and prepared following the manufacturer’s instructions. Prepared media included Blood Agar (BA), Mannitol Salt Agar (MSA), MacConkey Agar (MA), and Muller Hinton Agar (MHA). All biochemical test media were produced per Oxoid, UK, manufacturer specifications, and each swab sample was streaked onto both blood and mannitol salt agar plates to promote the growth of a wide range of bacterial species. The plates were then incubated aerobically at 37°C for 48 h. After incubation, colony morphology, and Gram staining were used to identify the different bacterial species present in the samples. Further biochemical tests, including catalase, oxidase, and sugar fermentation tests, were conducted to confirm the bacterial identification and sorting.

### Antibiotic Susceptibility Testing

The modified Kirby Bauer disk diffusion technique was employed. Two or three colonies were taken from each isolate and emulsified in 5 ml of sterile peptone water to achieve a turbidity of 0.5 McFarland. The prepared inoculum was applied to the MHA using a sterile inoculating loop. Penicillin G (1 g), gentamycin (10 g), clindamycin (2 g), erythromycin (15 g), tetracycline (30 g), trimethoprim-sulfamethoxazole (25 g), linezolid (10 g), vancomycin (5 g), and cefoxitin (30 g) antibiotic discs were utilized on the MHA. The reference strain of MRSA was ATCC 29523, obtained from the American Type Culture Collection. For 24 hours, the plates were kept at 37 degrees Celsius. Vernier calipers were used to measure the sizes of the inhibitory zones in millimeters (mm). The antibiotic discs were used to test their efficacy against the bacterial isolates, and the average diameter of the zone of inhibition was calculated. The obtained results were compared to cutoffs established by the Clinical and Laboratory Standards Institute (CLSI, 2017).

### Statistical Analysis

Data obtained from the antimicrobial susceptibility testing were analyzed using Microsoft Excel to assess the significance of differences between the bacterial profiles and susceptibility patterns of diabetic and non-diabetic controls. Descriptive statistics were used to summarize the data, and inferential statistics, such as t-tests or chi-square tests, were applied as appropriate. The results were then presented in tables and graphs to facilitate interpretation and discussion of the findings.

## Data Availability

All data produced in the present work are contained in the manuscript

## Supplementary Figures

**SF1:**
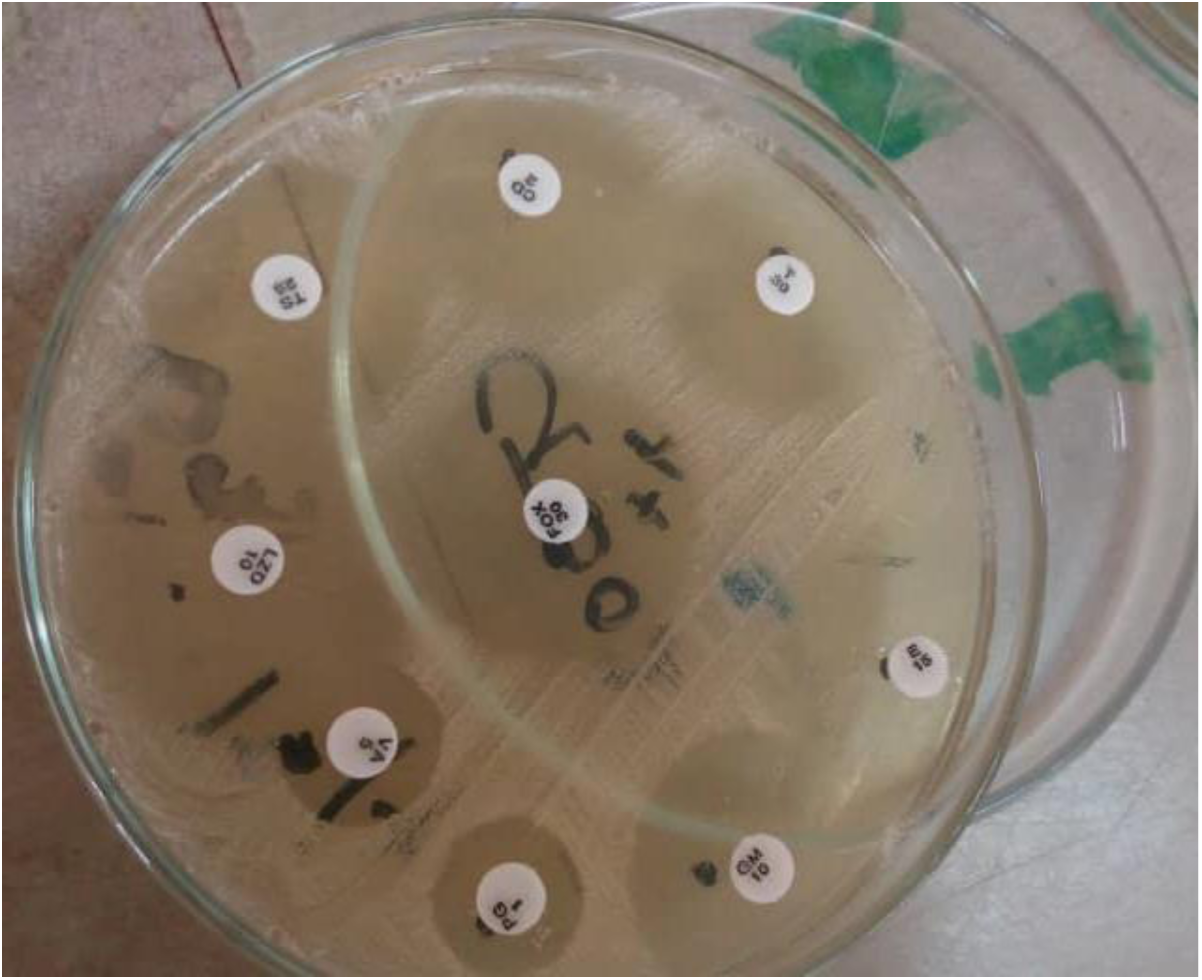
The image of Antibiotic susceptibility testing of S. aureus.

**SF2:**
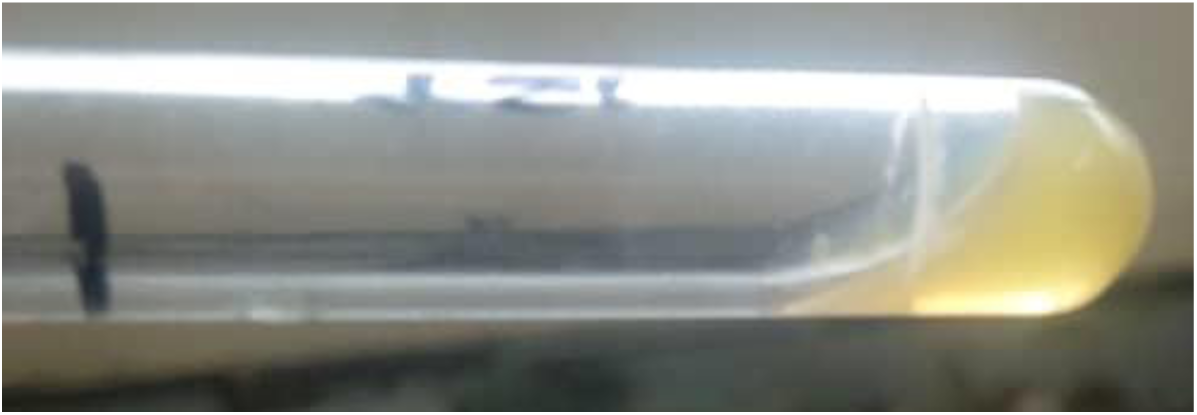
Positive Coagulase test for *S. aureus*

**SF3:**
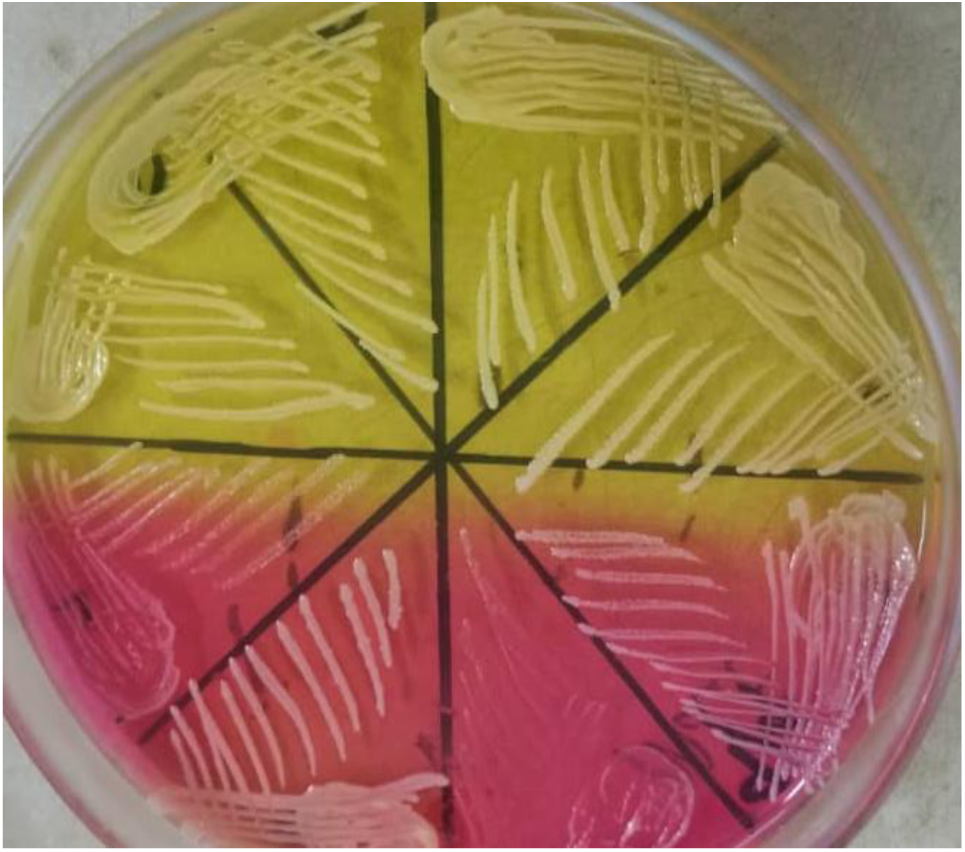
Confirmation of *S. aureus* on Mannitol salt agar

**SF4:**
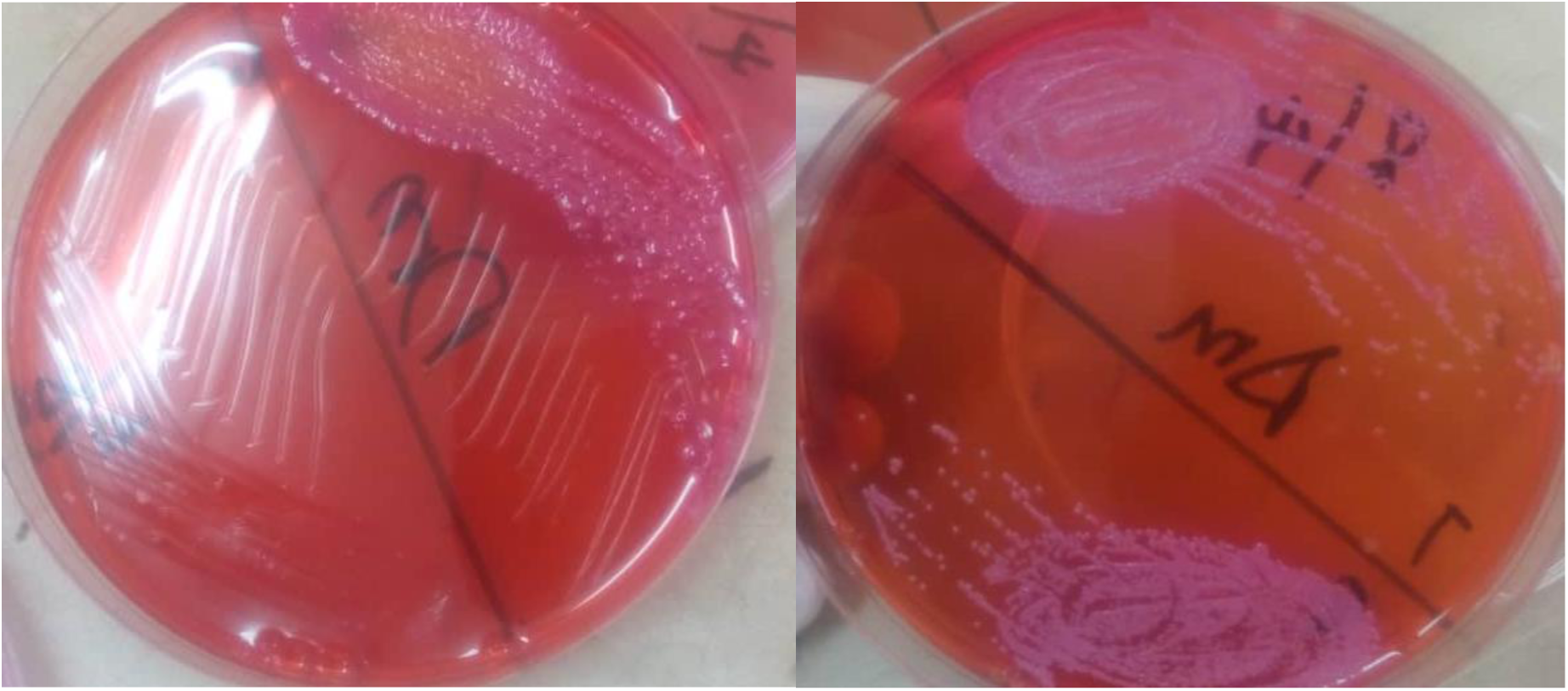
Bacteria growth from direct inoculation of swab stick samples onto Mackonkey agar

